# ACUTE BLOOD PRESSURE RESPONSES TO DIFFERENT RESISTANCE EXERCISES USING HIGH- AND LOW-INTENSITY VELOCITY-BASED TRAINING PROTOCOLS IN INDIVIDUALS WITH HYPERTENSION

**DOI:** 10.1101/2025.03.24.25324531

**Authors:** Luis A. Benavides-Roca, Germán Parra, Antonio R. Zamunér

## Abstract

Hypertension is a chronic condition that requires proper management to prevent cardiovascular complications, and resistance exercise is a recommended strategy for its control. This study aimed to determine the acute effects of specific exercises, commonly used in resistance training programs, performed at two intensities (low and high) with equal volume, on blood pressure in individuals with pharmacologically controlled hypertension. A crossover clinical trial was conducted with 26 participants diagnosed with hypertension, each completing two resistance training sessions one low-intensity and one high-intensity including squats, rows, deadlifts, and bench presses. The order of exercise intensity for the first session was randomly assigned, and blood pressure was measured using an automated device before and immediately after each exercise. Comparisons between baseline and post-exercise values were analyzed, with a significance level set at 5%. The results showed that systolic blood pressure (SBP) increased during the squat, rows, and deadlift compared to baseline, with the deadlift producing the highest values. For diastolic blood pressure (DBP), no significant differences were observed from baseline, but the squat and deadlift elicited higher values than the rows and bench press. Additionally, high-intensity training resulted in lower DBP values compared to the low-intensity protocol. In conclusion, resistance exercise increases SBP regardless of intensity, with the squat and deadlift producing the most significant changes, while high-intensity training leads to lower DBP values than low-intensity protocols.

## INTRODUCTION

Exercise is considered as one of the main non-pharmacological therapies for patients with hypertension [1]. Therefore, different modalities of exercise have been studied, including resistance training. Systematic reviews [2, 3] have shown that resistance exercise contributes to the reduction of blood pressure in stages of prehypertension and hypertension. Furthermore, post-exercise hypotensive effect that lasts over time has been observed [4, 5], especially when exercise is of moderate intensity [3]. Mechanisms such as nitric oxide release and decreased adrenergic discharge, which result in lower peripheral vascular resistance [6], have been proposed as contributing factors. However, studies evaluating the acute effects of specific resistance exercises at different intensities are incipient [7, 8]. There is evidence that resistance exercise produces an increase in blood pressure immediately after performance [7, 8] show an increase of approximately 20 mmHg in systolic (SBP) and 10 mmHg in diastolic blood pressure (DBP) with exercises such as inverted rows, squats, push-ups and sit-ups. Similarly, in hypertensive subjects, exercise of resistance at 80% intensity has been shown to increase SBP by approximately 6 mmHg, which is higher than the increase observed at 40% intensity [9].

Enhancing knowledge of the immediate effects of specific exercises on blood pressure could be useful for patients and therapists to identify exercises that may elicit higher increases in blood pressure in this population. Therefore, the aim of this study was to determine the acute effects of specific exercises, commonly used in resistance training programs (i.e. squats, rows, deadlift and bench-press), performed at two intensities (low and high intensity) with equal volume on blood pressure in individuals whit pharmacologically controlled hypertension.

## MATERIALS AND METHODS

### Study design

The present study is a secondary analysis of a randomized, single-blind, crossover clinical trial, registered at ClinicalTrials.gov (NCT06370546).

### Participants

Twenty-six participants of both sexes, aged between 30 and 60 years and with medical diagnosis of hypertension performed by a cardiologist according to the 2023 ESH guidelines [10], took part in the study. All the participants were undergoing pharmacological treatment.

Participants were excluded if they have any neurological, musculoskeletal or cardiovascular disease other than hypertension (e.g. atrial fibrillation, arrhythmias, and coronary disorders, pacemakers). In addition, participants were not included if they presented any contraindication for resistance exercise, were unable to perform the prescribed exercises, or reported to be drug abusers.

The study was performed in accordance with the Declaration of Helsinki, was approved by the local institutional ethics committee (omitted for review purposes) and all participants signed an informed consent.

**Fig 1.**
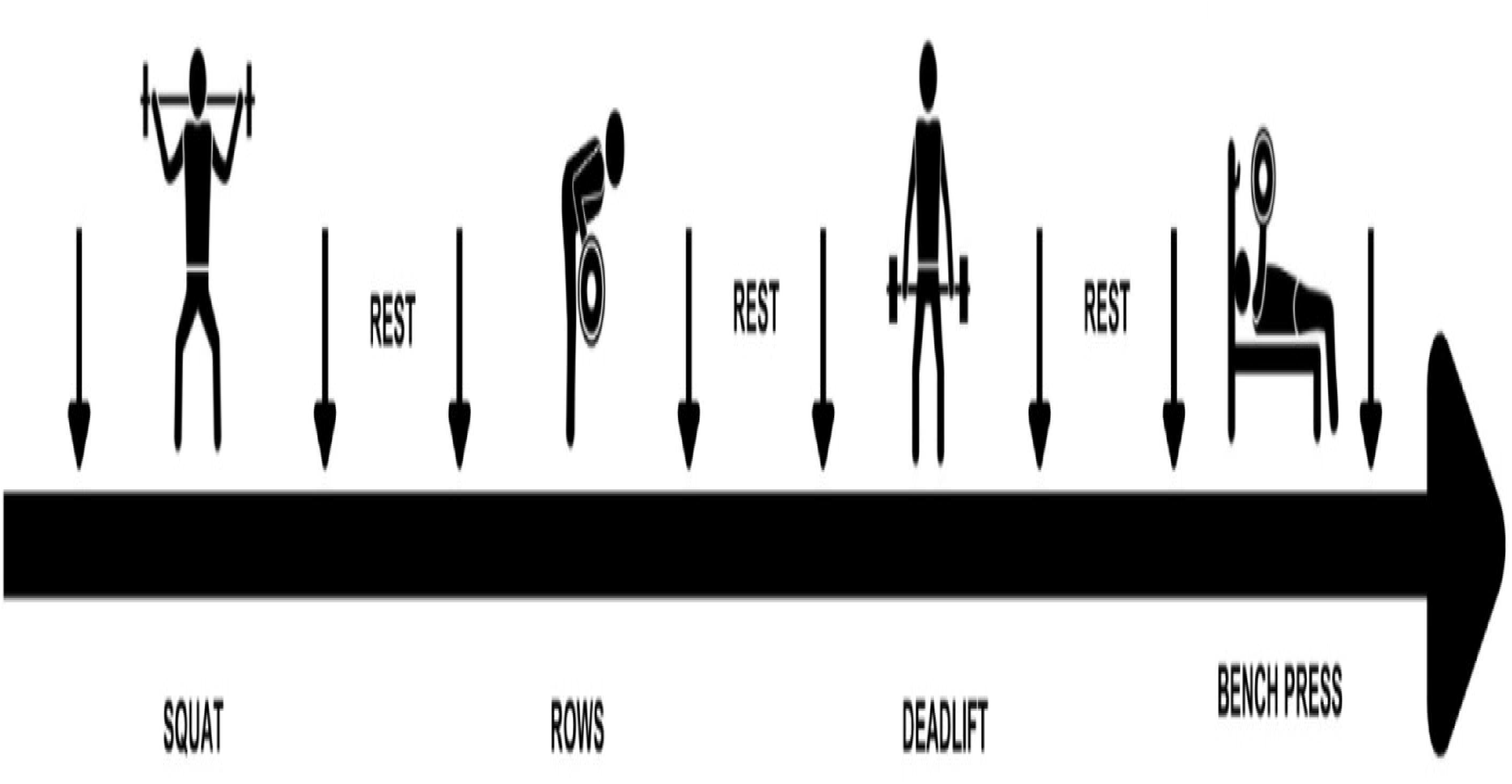
Procedure timeline. Arrows indicate blood pressure assessments.

### Procedure

All participants attended the Laboratory XXXX at the University XXXX (omitted for reviewing purpose) in three opportunities. In the first visit inclusion and exclusion criteria were reviewed, participants signed an informed consent. A full anamnesis was performed, anthropometric characteristics were measured, and the participants were acquainted with the experimental protocol. Then, participants were requested to return in two other opportunities to perform the resistance exercise sessions.

### Randomization and blinding

In the second visit, a randomization was performed to determine the intensity of the intervention (low-intensity or high-intensity resistance training). After a 1-week washout period, participants first allocated to perform the low-intensity protocol returned to the laboratory to perform the high-intensity protocol and vice versa.

### Resistance training

Two resistance velocity-based training protocols, with low- and high-intensity, were conducted using free weights. The low-intensity session comprised 6 sets of 12 repetitions performed with at an intensity of 40% of one-repetition maximum (1RM, faster velocity of execution). In contrast, the high-intensity session consisted of 6 sets of 6 repetitions at 80% of 1RM (slower velocity of execution). The protocols were designed to equalize the total volume of the resistance training sessions [11]. The load required to reach the prescribed intensity was individually determined based on the velocity of execution, as proposed by Hernandez-Belmonte et al. [13] and Benavides-Ubric et al. [14]. Therefore, participants were instructed to perform each repetition at their maximal intended velocity, which was continuously monitored in real time using a previously validated linear transducer (ADR®, Castilla, Spain) [15].

Before the intervention, a 7–10-minute warm-up was conducted, consisting of joint movements for the upper and lower body, aligned with the exercises included in the resistance training. During the warm-up, execution speed for each exercise was measured as part of the approximation series. This evaluation allowed for the determination of the appropriate weights for each participant, tailored to the required session intensity.

The exercises were 1) squats: consisting of a triple flexion of the lower limb (ankle, hip, and knee) with external resistance placed on the back using a barbell (execution velocity: low intensity = 1.15±0.09 m/s; high intensity = 0.63±0.05 m/s); 2) Reclined rows: participants were standing maintaining the trunk flexion. Participants were instructed to bringing the barbell close to the abdomen with elbow flexion and shoulder extension (execution velocity: low intensity = 1.42±0.10 m/s; high intensity = 1.23±0.07 m/s); 3) Deadlift: Participants lifted a bar from the ground while maintaining the core contraction and performing a trunk, knee and hip extensions (execution velocity: low intensity = 1.02±0.09 m/s; high intensity = 0.57±0.05 m/s); and 4) Bench press: participants were instructed to stay in supine posture on a bench, with both feet on the ground and to raise a barbell by flexing the shoulders and extending the elbows (execution velocity: low intensity = 1.14±0.09 m/s; high intensity = 0.50±0.06m/s).

After each exercise, participants rested until they felt fully recovered, and hemodynamic variables returned to baseline values (approximately 10 minutes).

### Main outcomes

The main outcomes measured were blood pressure, which was recorded before the warm-up, after a prolonged rest of 10 minutes in position supine, and after completing the series of each exercise.

The measurements included SBP and DBP, taken using a digital automated device (Omron HEM-907 sphygmomanometer, Healthcare, Tokyo, Japan).

### Statistical Analysis

The SPSS Statistics version 22 (New York, IBM Corp) program using the IBM Statics for Windows. The Shapiro-Wilk test was applied to check the normality of the data. A two-way repeated measures analysis of variance (ANOVA) was performed to assess interaction between intensity (high *vs* low) and time (baseline *vs* exercise). A significance value of p < 0.05 was considered for the analysis of both comparisons.

## RESULTS

Table 1 shows the demographic characteristics of the participants.

**Table 1.**
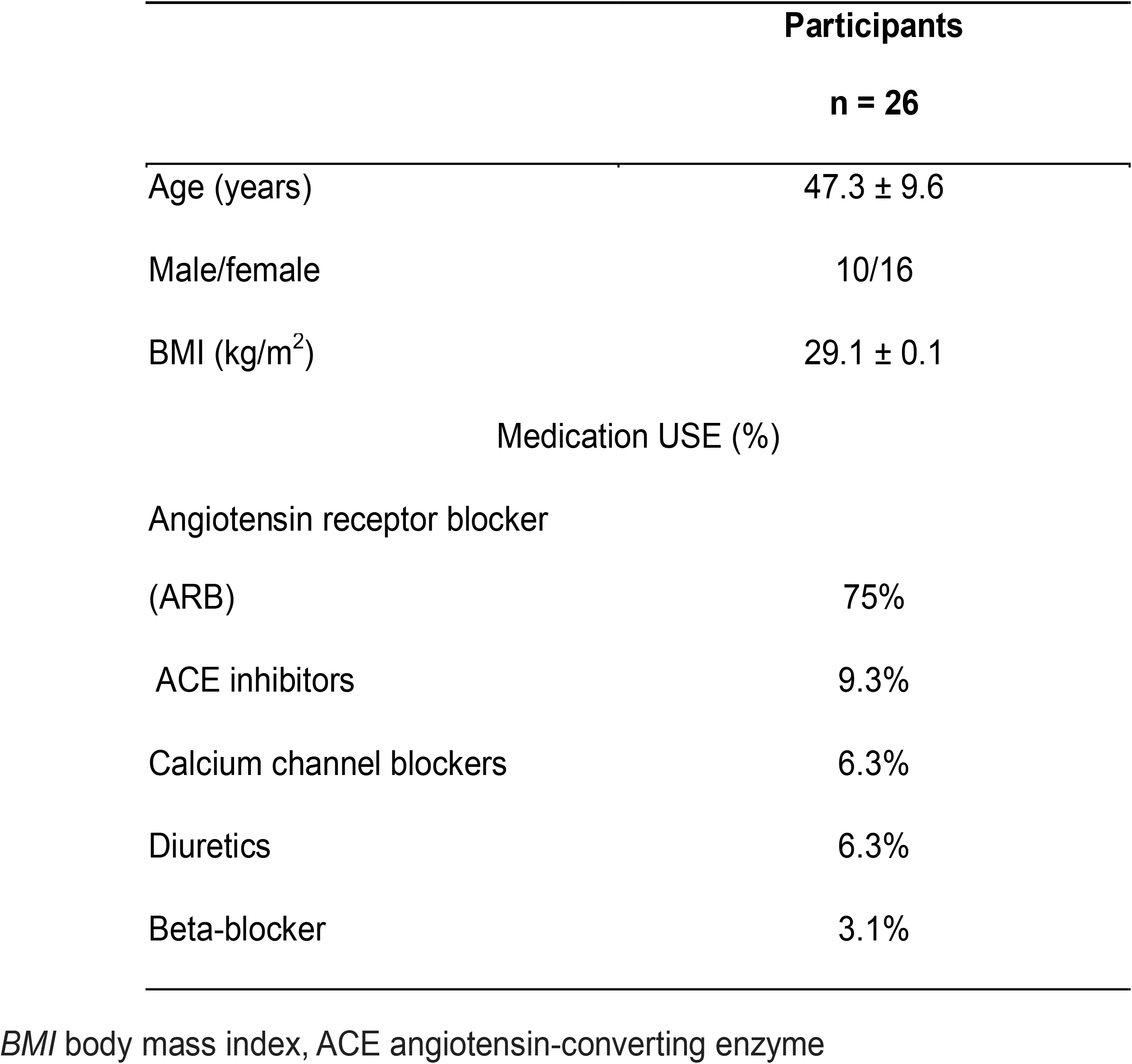
Demographic characteristics of participants.

Table 2 presents the blood pressure values for each exercise and the intensity at which they were performed. The analysis showed no significant interaction between intensity and exercise for SBP (F_4,96_ = 1.35; p = 0.25) and DBP (F_4,96_ = 1.9; p = 0.12). However, a significant main effect of exercise was observed for all outcomes (SBP and DBP [p < 0.001]).

**Table 2.** blood pressure assessed pre and immediately after the exercise resistance training.

|  |  | High-Intensity | Low-Intensity | *p-values* |  |  |
| --- | --- | --- | --- | --- | --- | --- |
|  |  |  |  | *Intensity* | *Exercise type* | *Interaction (E\*I)* |
| *SBP (mmHg)* | Baseline | 121±11.7 | 124±10.1 |  |  |  |
|  | Squat <sup>a,c</sup> | 134±17.9 | 138±15.4 |  |  |  |
|  | Rows <sup>a,b,c</sup> | 132±14.3 | 132±11.5 | 0.60 | <0.0001 | 0.25 |
|  | Deadlift <sup>a</sup> | 138±15 | 137±10.9 |  |  |  |
|  | Bench Press <sup>b</sup> | 126±14.6 | 127±9.9 |  |  |  |
| *DBP (mmHg)* | Baseline | 81±7.7 | 83±9.0 |  |  |  |
|  | Squat <sup>c,d</sup> | 83±8.9<sup>cf</sup> | 86±8.5<sup>cf</sup> |  |  |  |
|  | Rows | 77±9.7<sup>d</sup> | 83±7.7<sup>f</sup> | 0.003 | <0.0001 | 0.12 |
|  | Deadlift <sup>c,d</sup> | 83±8.1<sup>f</sup> | 86±6.5<sup>f</sup> |  |  |  |
|  | Bench Press | 78±7.8 | 79±6.1 |  |  |  |

<sup>a</sup> p<0.05 vs Baseline; <sup>b</sup> p<0.05 vs Deadlift; <sup>c</sup> p<0.05 vs Bench press; <sup>d</sup> p<0.05 vs Rows. 2-way ANOVA results for the interaction between Exercise Type and Intensity

Regardless the intensity, the data for SBP indicate that the baseline values were lower than those recorded during the squat (p < 0.001), rows (p < 0.001), and deadlift (p <0.001). Additionally, the bench press yielded lower values compared to the squat (p <0.001), rows (p = 0.01), and deadlift (p < 0.001). Furthermore, the deadlift produced higher SBP than the rows (p = 0.01). DBP data indicate that regardless of intensity, none of the exercises showed significant differences compared to baseline. However, the squat elicited higher values than the rows (p = 0.001) and the bench press (p <0.001). Similarly, the deadlift exhibited higher pressure values compared to the rows (p = 0.001) and the bench press (p <0.001). In addition, a significant main effect of intensity, was found for DBP (p=0.003), indicating that high-intensity training resulted in lower values compared to the low-intensity protocol.

To account for a possible effect of sex, two- and three-way ANOVAs were performed to assess the interactions between intensity, exercise, and sex. No significant three-way interaction (exercise × intensity × sex) was found (SBP: F_4,96_ = 1.1, p = 0.37 ; DBP: F_4,96_ = 1.2, p = 0.28), nor were the two-way interactions exercise × sex (SBP: F_4,96_ = 1.0, p = 0.39; DBP: F_4,96_ = 0.5, p = 0.71) or intensity × sex (SBP: F = 0.23, p = 0.64; DBP: F = 0.18, p = 0.68).

## DISCUSSION

The purpose of this study was to determine the acute effects on blood pressure of specific exercises commonly used in resistance training programs, performed at two intensities (low and high intensity). The main findings revealed that 1) squats and deadlift were the exercises eliciting the highest SBP and DBP, regardless the intensity; and 2) DBP was lower after the high intensity exercises compared to the low intensity protocol.

These findings are in agreement with the results reported by Bond et al. [7], who evaluated subjects with prehypertension during handgrip exercises. The observed cardiovascular effects are consistent with the scientific statement from the American Heart Association [15], which highlights that during both dynamic and isometric exercise in healthy individuals, SBP tends to increase due to a rise in cardiac output, while DBP either remains stable or decreases slightly. This response is attributed to the interplay between cardiac output and peripheral vascular resistance, which collectively regulate blood pressure [16].

These results are important to characterize the blood pressure response immediately after exercise commonly used in clinical settings or in the context of cardiovascular rehabilitation. The hypertensive response to the resistance training observed in this study is in accordance with previous studies [17, 18]. It is well established that physical exercise increases heart rate and stroke volume to meet the demands of the muscles. This is supported by the research of Vale et al. [18], which observed a significant rise in SBP immediately after exercise with loads of 6 RM (high intensity) and 15 RM (low intensity). This SBP response was also observed in older women with hypertension, whit SBP increased by approximately 10 mmHg following exercises such of bench press, leg press, and lat pull-downs (3 sets of 10 repetitions) [19].

Based on the above, the results obtained may be influenced by the planning of the strength training session, as the equalization of training volume could be linked to the lack of significant differences between the high-intensity and low-intensity groups after each exercise. These findings highlight the importance of training volume as a crucial factor in strength training programming [20]. Effective volume management is essential for eliciting specific neuromuscular adaptations [21] and inducing blood pressure adjustments in hypertensive individuals [22]. Previous research has shown that similar neuromuscular adaptations can occur at different training intensities when the workload is equalized between groups [23]. These results are consistent with those observed in the present study, where training volume was equal across groups. Furthermore, longitudinal studies have demonstrated statistically significant reductions in blood pressure in sedentary patients using low training volumes (two sets per muscle group and exercise). For instance, a decrease in mean SBP from 152 mmHg to 122 mmHg and DBP from 83 mmHg to 73 mmHg was observed after 24 sessions over 12 weeks [22]. This suggests that training volume is a modifiable variable that can induce cardiovascular adaptations.

For DBP, the response observed differs from that of SBP, as the results show no significant difference from baseline. This is consistent with the findings of Stöhr et al. [24], who also reported no significant changes in diastolic parameters (such as end-diastolic volume and early and late peak diastolic blood velocity) after a leg press session at both high (60% of repetition maximum) and low intensity (30% of repetition maximum). However, the literature presents mixed results, Marques et al. [25] observed an increase in DBP with both high (6.8% increase) and low (5.9% increase) training volume strength stimuli in older adults (78.9 ± 7.2 years old). Conversely, decreases in DBP (−18 mmHg) have been observed in individuals with hypertension and diabetes following strength training at 75% of one repetition maximum [26]. Similarly, a reduction in DBP during the 15 minutes after self-loaded exercise (5.7 ± 1.5 mmHg) has been reported in physically active hypertensive individuals [8]. Therefore, it is imperative to analyze DBP results in relation to the type of stimulus applied and the characteristics of the load used. These factors have the capacity to reduce vascular resistance and induce vasodilation, which are key mechanisms influencing DBP [27]. This may provide a rationale for the higher DBP values observed in low-intensity exercise compared to high-intensity exercise. This finding is consistent with the investigation of De Souza et al. [28], who reported that hypertensive subjects exhibited higher blood pressure values (+2 mmHg) when exercising at 40% of their one-repetition maximum compared to 80%.

Another worth noting finding of the present study was that the deadlift and squat exercises were the exercise who increased the SBP the most, regardless of the training session’s intensity. These exercises engage a significant amount of muscle mass and are performed within a closed kinetic chain. Their execution activates the lower-body muscles and generates complementary contractions that regulate movement and amplify the pressor response due to the large number of muscles involved [29]. Furthermore, the mechanical characteristics of these exercises— particularly the involvement of hip flexion and extension—have been shown to increase cardiac output because of orthostatic stress [30]. Therefore, from a clinical perspective, it is important that therapists consider the baseline blood pressure values before every resistance training session, especially when prescribing squat and deadlift exercises.

### Limitation

Despite the results, some limitations must be pointed out. In the current study was not possible to measure blood pressure continuously during movement execution, due to technical limitations of the device used. This measurement would have allowed us to analyze the blood pressure response to different phases of muscle contraction and to determine the peak for each exercise, which also could be useful for patients and therapists to identify the important moments of rise and fall in blood pressure. Therefore, future studies should consider continuously monitoring the non-invasive blood pressure during resistance training sessions.

## CONCLUSIONS

In conclusion, squats and deadlifts performed using a velocity-based protocol elicited the highest systolic and diastolic blood pressure responses, regardless of intensity. Additionally, diastolic blood pressure was lower following high-intensity exercises compared to the low-intensity protocols. Future studies should investigate the chronic effects of velocity-based resistance exercises on blood pressure of individuals with hypertension.

## ABREVIATURES

SPD: Systolic blood pressure
DBP: Diastolic blood pressure

## Data Availability Statement

The data this article will be shared on reasonable request to the corresponding author.

## AUTHOR CONTRIBUTIONS

LB, AZ and GP contributed to the conception and design. LB, AZ and GP contributed to data acquisition and data interpretation of the work. LB, AZ and GP drafted the manuscript. AZ and GP critically revised the manuscript.

## ETHICS

This study was approved by the ethics committee of the University Catholic of Maule under protocol #248/2022.

## COMPETING INTERESTS

The authors declare no competing interests.

## Notes

**Conflict of interest:** none declared

### Competing Interest Statement

The authors have declared no competing interest.

### Clinical Trial

NCT06370546

### Funding Statement

This study did not receive any funding

### Author Declarations

The Ethics committee of University Catholic of Maule approval for this work

## REFERENCE

1. Lopes S, Mesquita-Bastos J, Garcia C, Bertoquini S, Ribau V, Teixeira M, et al. Effect of exercise training on ambulatory blood pressure among patients with resistant hypertension: a randomized clinical trial. JAMA Cardiol. 2021;6(11):1317–1323.

2. Henkin JS, Pinto RS, Machado CL, Wilhelm EN. Chronic effect of resistance training on blood pressure in older adults with prehypertension and hypertension: a systematic review and meta-analysis. Exp Gerontol. 2023;177:112193.

3. Oliver-Martinez PA, Ramos-Campo DJ, Martinez-Aranda LM, Martinez-Rodriguez A, Rubio-Arias JA. Chronic effects and optimal dosage of strength training on SBP and DBP: a systematic review with meta-analysis. J Hypertens. 2020;38(10):1909–1918.

4. Correia RR, Veras ASC, Tebar WR, Rufino JC, Batista VRG, Teixeira GR. Strength training for arterial hypertension treatment: a systematic review and meta-analysis of randomized clinical trials. Sci Rep. 2023;13(1):201.

5. Nascimento DDC, da Silva CR, Valduga R, Saraiva B, de Sousa Neto IV, Vieira A, et al. Blood pressure response to resistance training in hypertensive and normotensive older women. Clin Interv Aging. 2018;541–553.

6. Otsuki T, Nakamura F, Zempo-Miyaki A. Nitric oxide and decreases in resistance exercise blood pressure with aerobic exercise training in older individuals. Front Physiol. 2019;10:1204.

7. Bond V, Curry BH, Adams RG, Obisesan T, Pemminati S, Gorantla VR, et al. Cardiovascular responses to an isometric handgrip exercise in females with prehypertension. N Am J Med Sci. 2016;8(6):243.

8. Ferrari R, Cadore EL, Perico B, Kothe GB. Acute effects of body-weight resistance exercises on blood pressure and glycemia in middle-aged adults with hypertension. Clin Exp Hypertens. 2021;43(1):63–68.

9. Cavalcante PAM, Rica RL, Evangelista AL, Serra AJ, Figueira Jr A, Pontes Jr FL, et al. Effects of exercise intensity on postexercise hypotension after resistance training session in overweight hypertensive patients. Clin Interv Aging. 2015;1487–1495.

10. Mancia-Chairperson G, Brunström M, Burnier M, Grassi G, Januszewicz A, Muiesan ML, et al. 2023 ESH Guidelines for the management of arterial hypertension: The Task Force for the management of arterial hypertension of the European Society of Hypertension Endorsed by the European Renal Association (ERA) and the International Society of Hypertension (ISH). J Hypertens. 2023;41(12):1874–2071.

11. Rodriguez-Lopez C, Alcazar J, Sánchez-Martín C, Ara I, Csapo R, Alegre LM. Mechanical characteristics of heavy vs. light load ballistic resistance training in older adults. J Strength Cond Res. 2022;36(8):2094–2101.

12. Hernández-Belmonte A, Buendía-Romero Á, Pallares JG, Martínez-Cava A. Velocity-based method in free-weight and machine-based training modalities: The degree of freedom matters. J Strength Cond Res. 2023;37(9):e500–e509.

13. Benavides-Ubric A, Díez-Fernández DM, Rodríguez-Pérez MA, Ortega-Becerra M, Pareja-Blanco F. Analysis of the load-velocity relationship in deadlift exercise. J Sports Sci Med. 2020;19(3):452.

14. Lopez-Torres O, Fernandez-Elias VE, Li J, Gomez-Ruano MA, Guadalupe-Grau A. Validity and reliability of a new low-cost linear position transducer to measure mean propulsive velocity: The ADR device. Proc Inst Mech Eng P J Sports Eng Technol. 2022;17543371221104345.

15. Bittner VA, Coke LA, Fleg JL, Forman DE, Gerber TC, Gulati M, et al. Exercise standards for testing and training. Circulation. 2013;128:873–934.

16. Hu XQ, Zhang L. Oxidative regulation of vascular Cav1.2 channels triggers vascular dysfunction in hypertension-related disorders. Antioxidants (Basel). 2022;11(12):2432.

17. Galvão L, Póvoa TIR, Jardim PCV, Lima AL, Barroso WKS, Seguro CS, et al. Acute effects of high-intensity resistance training on central blood pressure parameters of elderly hypertensive women: A crossover clinical trial. J Hypertens. 2023;41(6):912–917.

18. Vale AF, Carneiro JA, Jardim PC, Jardim TV, Steele J, Fisher JP, et al. Acute effects of different resistance training loads on cardiac autonomic modulation in hypertensive postmenopausal women. J Transl Med. 2018;16:1–9.

19. Galvão L, Póvoa TIR, Jardim PCV, Lima AL, Barroso WKS, Seguro CS, et al. Acute effects of high-intensity resistance training on central blood pressure parameters of elderly hypertensive women: A crossover clinical trial. J Hypertens. 2023;41(6):912–917.

20. Paulsen GO, Myklestad D, Raastad T. The influence of volume of exercise on early adaptations to strength training. J Strength Cond Res. 2003;17(1):115–120.

21. Fonseca FS, Costa BDDV, Ferreira MEC, Paes S, de Lima-Junior D, Kassiano W, et al. Acute effects of equated volume-load resistance training leading to muscular failure versus non-failure on neuromuscular performance. J Exerc Sci Fit. 2020;18(2):94–100.

22. Seguro CS, Rebelo ACS, Silva AG, Dos Santos MMA, Cardoso JS, Apolinário V, et al. Use of low volume, high effort resistance training to manage blood pressure in hypertensive patients inside a public hospital: a proof of concept study. Eur J Transl Myol. 2021;31(1).

23. Radaelli R, Botton CE, Wilhelm EN, Bottaro M, Lacerda F, Gaya A, et al. Low- and high-volume strength training induces similar neuromuscular improvements in muscle quality in elderly women. Exp Gerontol. 2013;48(8):710–716.

24. Stöhr EJ, Stembridge M, Shave R, Samuel TJ, Stone KJ, Esformes JI. Systolic and diastolic LV mechanics during and following resistance exercise. Med Sci Sports Exerc. 2017;49(10):2025–2031.

25. Marques DL, Neiva HP, Fail LB, Gil MH, Marques MC. Acute effects of low and high-volume resistance training on hemodynamic, metabolic and neuromuscular parameters in older adults. Exp Gerontol. 2019;125:110685.

26. De Sousa RAL, Hagenbeck KF, Arsa G, Pardono E. Moderate/high resistance exercise is better to reduce blood glucose and blood pressure in middle-aged diabetic subjects. Rev Bras Educ Fis Esporte. 2020;34(1):165–175.

27. Sushma T, Sangeeta G, Tiwari SK, Girish S. Effect of isotonic exercise (walking) on various physiological parameters in hypertension. J Stress Physiol Biochem. 2011;7(3):122–131.

28. De Souza Nery S, Gomides RS, da Silva GV, de Moraes Forjaz CL, Mion Jr D, Tinucci T. Intra-arterial blood pressure response in hypertensive subjects during low-and high-intensity resistance exercise. Clinics (Sao Paulo). 2010;65(3):271–277.

29. Mayo JJ, Kravitz LEN. A review of the acute cardiovascular responses to resistance exercise of healthy young and older adults. J Strength Cond Res. 1999;13(1):90–96.

30. Chukwuemeka UM, Benjamin CP, Uchenwoke CI, Okonkwo UP, Anakor AC, Ede SS, et al. Impact of squatting on selected cardiovascular parameters among college students. Sci Rep. 2024;14(1):5669.

